# Effects of physical activity and exercise on well-being in the context of the Covid-19 pandemic

**DOI:** 10.1101/2020.06.08.20125575

**Authors:** Juliana Marques de Abreu, Roberta Andrade de Souza, Livia Gomes Viana-Meireles, J. Landeira-Fernandez, Alberto Filgueiras

## Abstract

**Background:** A disease discovered in China, COVID-19, was characterized by the World Health Organization (WHO) as a pandemic in March 2020. Many countries in the world implemented social isolation as a strategy to contain the virus transmission. The same physical distancing which protects society from COVID-19 from spreading may have an impact on the mental health and well-being of the population This study aims to shed some light on this phenomenon by assessing the relationship between physical activity and SWB among individuals in the social isolation period of COVID-19.

**Methods:** Data were collected in Brazil between March 31^st^ and April 2^nd^, 2020. All volunteers agreed to participate by digitally checking the option of agreement right after reading the Consent Terms. The inclusion criteria were participants over 18 years old who had been in social isolation for at least one week and agreed to the Consent Terms. Three instruments were used: a questionnaire was built for this study which aimed to assess the participants’ exercise routine. The second instrument called Psychosocial Aspects, Well-being and Exercise in Confinement (PAWEC) was also created by these researchers and aimed to assess the relationship between well-being and physical activity during the social isolation period. And the third measure was the Brazilian Portuguese-adapted version of the Positive and Negative Affect Schedule (PANAS).

**Findings:** A total of 592 participants reported being in social isolation for an average of 14.4 (SD=3.3) days. The amount of participants who reported strength training as exercise increased from 31 (5.2%) before isolation to 82 (13.9%) during quarantine. The study shows that well-being related to the practice of physical activity during quarantine is linked to an established routine of physical activity prior to the social isolation period.

**Interpretation:** People who already practiced physical activity feel more motivated to continue practicing during this period and this causes the appearance of positive affects, unlike people who are only now starting to exercise; according to the study, negative aspects can occur for those who are only just starting. In a period of social isolation, it is important that the practice of physical activity is closer to previous habits, also finding that an obligation to exercise during this period when this was not a reality for the person can contribute to an increase in malaise.

## Introduction

The Corona Virus Disease 2019 (Covid-19) was discovered in China in December 2019 in the city of Wuhan, and soon became a public health emergency of international interest (Wang et al., 2020). Covid-19 is an infectious disease caused by the coronavirus of severe acute respiratory syndrome, which can cause respiratory infections. The clinical conditions vary from mild to moderate and at times evolve into severe. In its milder form, the manifestation of symptoms is similar to that of a common flu: symptoms such as fever, coughing, difficulty in breathing, muscle pain, headache and a sore throat and runny nose. The virus can be transmitted from one person to another through droplets of saliva or mucus, expelled through the mouth or nostrils when an infected person coughs or sneezes (WHO, 2020).

On March 11^th^, 2020, Covid-19 was characterized by the World Health Organization (WHO) as a pandemic. One of the preventive measures suggested by the WHO to contain its contagious spread was physical distancing between people, instructing everyone to stay in their homes (Findik et al., 2012; (Geelhoed, 1978); Ward, 2000; WHO, 2020). An interruption of daily activities such as work, studies, and leisure, among others, occurred in an attempt to prevent the increase in cases in the country. Since the beginning of March, most states in Brazil have adhered to social isolation and the population constantly refers to this period as quarantine (Brazil, 2020). Social isolation is the behavior where a person or a group of people, voluntarily or involuntarily, withdraws from social interactions and activities, therefore decreasing the chances of greater spread of diseases (Hawryluck et al., 2004).

In the COVID-19 pandemic, asymptomatic people may transmit the virus, so there is a great need for social distancing for the entire population, not only including those at risk or including who are proven to be infected. The Ministry of Health in Brazil published through ordinance 356 of March 11^th^, 2020, that “…isolation policies aim to separate symptomatic and asymptomatic people diagnosed with COVID-19 from the rest of the population in order to prevent the spread of infection and local transmission” (Brazil, 2020).

However, the same physical distance, which protects COVID-19 from spreading, may have an impact on mental health and the well-being of the population. In a 2004 study of 129 Canadian people who were quarantined due to the SARS epidemic outbreak, symptoms of post-traumatic and depressive stress were observed. The symptoms were directly related to age, educational level, living with other adults and not having children, and the duration of quarantine. The longer in isolation, the greater the risk of symptoms to appear (Hawryluck et al., 2004). The same results were found in similar studies of SARS quarantine in Taiwan (Bai et al., 2004) and Hong Kong (Lee et al., 2005). Filgueiras and Stults-Kolehmainen (2020) recently studied the psychosocial factors among Brazilians in quarantine due to Covid-19 and found that gender, quality of nutrition, attendance in tele-psychotherapy, exercise frequency, presence of older adults in quarantine with the person, obligation to work outside, education level (more educated, lesser risk for mental illness) and age (younger age, greater risk) predicted depression and anxiety states.

According to the most consensual and empirically tested definition, Subjective Well-Being (SWB) refers to understanding how people assess their own lives. Such assessments must be cognitive (overall satisfaction with life and other specific domains such as marriage and work) and must also include a personal analysis of the frequency with which positive and negative emotions are experienced. For an adequate level of SWB to be reported, it is necessary for the individual to recognize higher levels of life satisfaction, a high frequency of positive emotional experiences and a low frequency of negative emotions (Siqueira & Padovam, 2008). This leads to the assumption that lower levels of SWB are also linked to higher levels of psychosocial symptoms such as anxiety, depression and stress (Whitehead et al., 2019). In fact, there is evidence that supports this hypothesis among different types of samples, such as: children and adolescents (Pate et al., 2019), young adults (Whitehead et al., 2019) and older adults (Cheng et al., 2019).

Recent evidence supports the pivotal role of physical activity and exercise to decrease stress, depression and anxiety-like symptoms (Cid et al., 2007; Berger, 2004; Stuart & Nanette, 2007; Buckworth & Dishman, 2002; Kekäläinen et al., 2019; Panza et al., 2019; (Singleton, 2019). The International Society of Sport Psychology (Singer, 1992), provided a consensus statement linking physical activity and psychological benefits, and their conclusion was that long-term exercise is generally associated with reduced anxiety and stress levels, a decrease in depression levels, and an increase in subjects’ self-esteem and positive emotions.

According to the meta-analysis carried out by (Stuart & Nanette, 2007), sessions of twenty to sixty minutes, three to five times a week, with an intensity between 60%-90% of the maximum cardiac frequency (FCMax) are the key factors for physical exercise to generate more consistent psychological benefits (Berger, 2004; Buckworth & Dishman, 2002; A.Cruz, 1996; Samulski & Noce, 2000). Thus, the main hypothesis based on this evidence provided by the literature so far is that physical activity leads to reduced psychosocial symptoms and negative affect, whereas it also means increase in positive affect and overall SWB. However, it is unclear whether people in quarantine are able to engage in such frequency and regularity of exercise and if this practice is enough to provide SWB. The present research aims to shed some light on this phenomenon by assessing the relationship between physical activity and SWB among individuals in the social isolation period by COVID-19.

## Method

### Sample

The sample consisted of a total of 592 participants [371 women (62.7%), 220 men (37.2%) and 1 transgender (0.01%)], with an average age of 32.3 years (SD=10.5). All volunteers agreed to participate by digitally checking the option of agreement right after reading the Consent Terms.

### Procedures

This study was conducted between March 31^st^ and April 2^nd^, 2020. The project was approved by the Ethical Committee of the last author’s institution under the protocol no.2020.876-459. All procedures were in accordance with the Helsinki Declaration and the Ethical guidelines of the Brazilian authorities. Volunteers were recruited via authors’ social media and messaging smartphone apps. Upon receiving the link to answer the questionnaire, the participant had access to the research presentation and Consent Terms, explaining that this research was voluntary and non-mandatory and that the information obtained would be kept anonymous. After agreement to participate, those who did not accept were directed to a thank you page; whereas the others were addressed to the demographic questionnaire. The appearance order of questions and instruments were always the same: (1) Consent Terms, (2) demographic questionnaire, (3) exercise routine, (4) PAWEC, (5) PANAS, (6) Thank you page.

The inclusion criteria were participants over 18 years old who had been in social isolation for at least one week and agreed to the Consent Terms. Exclusion criteria were volunteers with a history of any kind of psychiatric condition, even those under treatment, and those who self-reported to be sedentary.

### Instruments

Three instruments were used online and sent in a single form of *Google Docs*. Respondents first had access to the Consent Terms. After agreement to participate, the sociodemographic section was presented (age, gender, education, number of days in quarantine, physical activity and exercise habits) followed by the three questionnaires in separate sections.

The first questionnaire was created for this study. It is an 8-item instrument which aimed to assess the participants’ exercise routine. Five questions were answered in dichotomous multiple-choice options: 1 - “Yes” or 2 - “No”. Items answered using the previous scale were: (i) “Have you been monitored by an online fitness coach during quarantine?”, (ii) “Were you monitored by a fitness coach before isolation?”, (iii) “Did you use any media source (i.e., YouTube, Social Media, videos, smartphone apps, *etc*.) to exercise before the quarantine?”, (iv) “Do you use any media source (i.e., YouTube, Social Media, videos, smartphone apps, *etc*.) to exercise during isolation?” and (v) “Are you doing exercise more frequently now than before the quarantine?”.

Another two items (i.e., “What physical activities do you do?” and “What physical activities did you use to do before confinement?”) were answered using the following options: “strength training”, “functional training”, “yoga/pilates”, “martial arts/fighting”, “walking/running”, “dancing/zumba”, “bicycle”, “swimming” and “other”; participants were instructed to consider the type of exercise they did most frequently. The final question was responded using an open-ended form in which volunteers had to provide information regarding “the number of days you practice exercise or physical activity before isolation.”

The second instrument called Psychosocial Aspects, Well-being and Exercise in Confinement (PAWEC) was also created by these researchers and aimed to assess the relationship between well-being and physical activity during the social isolation period. It is an 18-item questionnaire which seeks to investigate whether the frequency that physical activity and exercise had a positive or a negative influence on psychological and SWB aspects, namely mood, happiness, motivation, anxiety and sadness. The answers were provided in a 4-point Likert scale ranging from “Always” to “Never” (specifically, 1 - “Always”, 2 - “Sometimes”, 3 - “Rarely” and 4 - “Never”). Among the 18 items of this measure, 8 items are scored inversely (items 10, 11, 12, 13, 14, 16, 17 and 18). Examples of items are: item 7 - “Do you feel happy while exercising in quarantine?”, item 9 - “How often do you believe it is important to exercise during isolation?” and item 13 - “Do you feel anxious whenever you exercise during quarantine?”.

The third measure was the Brazilian Portuguese-adapted version of the Positive and Negative Affect Schedule (PANAS) by Pires et al. (2013). It is a 20-item instrument to be answered in a 5-point Likert-type scale (ranging from 1 - “totally disagree” to 5 - “totally agree”) which was originally developed by Watson, Clark and Tellegen (1988), and aims to measure Positive Affect (AP) and Negative Affect (NA), defined as general dimensions that describe the affective experience of individuals. High scores in NA reflect subjective displeasure and malaise, including emotions such as fear, nervousness and disturbance. High levels of PA implicate subjective pleasure and well-being, including emotions such as enthusiasm, inspiration and determination. The Brazilian version of PANAS presented a two-factor structure of significantly moderate negative correlation between factors (*r*=-0.42) and reliability of α=0.84 measured by Cronbach’s alpha, ranging from α=0.88 in the positive affect dimension to α=0.90 in the negative affect factor (Pires et al., 2013).

### Data Analyses

Descriptive statistics were calculated according to the nature of the measure: frequency and percentage were provided for categorical data, whereas arithmetic average and standard deviation (SD) were provided for continuous data. Cronbach’s alpha (α) was computed in order to investigate preliminary reliability of PAWEC. The preliminary validity was assessed using an Exploratory Factor Analysis (EFA) which was performed adopting the (Baglin, 2014) recommendation for ordinal variables. The procedure to determine the number of factors was the Parallel Analysis (PA) using the polychoric correlation matrix, the Unweighted Least Square (ULS) factor modeling was performed to assess factor retention and the Direct Oblimin rotation was adopted as an oblique method if needed due to expectancy of correlated factors, even though significant negative correlation was expected.

After ensuring enough reliability and validity of both measures developed for this research, a linear multiple regression (LMR) was separately computed for PA and NA PANAS scores as dependent variables. The stepwise method was adopted in those regressions. The first step was the PAWEC total score, demographic (i.e., age, gender, education and number of days in quarantine) and exercise routine variables were considered to predict the results of both PANAS factors. The second step of the regression comprised the PAWEC items independently. The significance level for variable inclusion in the LMR was *p*<0.05, the beta (*β*) coefficient revealed the strength of the association level between independent and predicted variables, *t*-test statistics (in addition to the *p*-value and effect size) were computed to assess whether one variable would be included in the LMR or not, and an ANOVA was provided to compare the LMR model to the null-hypothesis (i.e. the constant). The effect size was measured by the *f*^2^ statistics of the respective ANOVA considering the following interpretation: above 0.02 and below 0.15 is a small effect size, above 0.15 and above 0.35 is considered a moderate effect size, and above 0.35 is a large effect size. Descriptive statistics and LMR were computed in the free R software program using the psych package. EFA was performed in the FACTOR software program (Baglin, 2014) and effect sizes were calculated in the G*Power version 3.1.9.2 software program.

## Results

Participants reported in the demographic questionnaire being in an average of 14.4 (SD=3.3) days in social isolation with other 2.7 (SD=2.3) people living with them during this period. Volunteers showed diverse levels of education, with 150 (25.3%) answering to have completed elementary school, 161 (27.1%) had a high school degree, 172 (28.9%) were attending College, 28 (4.7%) had a Bachelor’s degree, 41 (8.1%) reported to have a Master’s degree and 35 (5.9%) a Ph.D. Regarding marital status, 343 (57.9%) participants were single and 189 (31.9%) were married. Finally, volunteers responded to have 2.1 (SD=2.6) children. In addressing the exercise routines of the participants, the data showed that they engaged in physical activities on average 4.5 (SD=1.2) days per week. Categorical variables of the exercise routine of participants are shown in Table 1.

**Table 1.** Descriptive statistics of the categorical variables of the exercise routine questionnaire.

| Question | Frequency |  |
| --- | --- | --- |
|  | Number | Percentage |
| *Were you monitored by a fitness coach before* |  |  |
| Yes | 495 | 83.6% |
| No | 97 | 16.4% |
| *Have you been monitored by an online fitness* |  |  |
| Yes | 252 | 42.6% |
| No | 340 | 57.4% |
| *Did you use any media source (i.e., YouTube,* |  |  |
| Yes | 143 | 24.2% |
| No | 449 | 75.8% |
| *Do you use any media source (i.e., YouTube,* |  |  |
| Yes | 352 | 59.5% |
| No | 240 | 40.5% |
| *Are you practicing exercise more frequently* |  |  |
| Yes | 101 | 17.1% |
| No | 491 | 82.9% |

Type of exercise before and during quarantine changed frequency, although it is not possible to infer statistical differences. The amount of participants who reported strength training as exercise increased from 31 (5.2%) before isolation to 82 (13.9%) during quarantine; functional training also increased from 26 (4.4%) to 292 (49.3%), yoga/pilates practice decreased from 63 (10.6%) to 36 (6.1%), martial arts/fighting also decreased with 107 (18.1%) of the volunteers reporting to practice it before quarantine and only 10 (1.7%) during isolation; walking/running increased as the main exercise for people from 27 (4.6%) to 71 (12.0%) before and during confinement, respectively; dancing/zumba remained almost the same, being reported to be the main exercise of 28 (4.7%) before quarantine to 27 (4.6%) during isolation; bicycle training (either exercise bike or traditional cycling) decreased from 162 (27.4%) to 19 (3.2%); swimming also decreased from 99 (16.7%) to 2 (0.3%), and other types of exercise including possible combinations increased from 49 (8.3%) before isolation to 53 (9.0%) during confinement.

The psychometric properties of PAWEC were to be provided before proceeding with other analyses. Preliminary EFA results showed sample adequacy for *KMO*=0.849 and a significant Bartlett sphericity test: χ^2^=3509.238, *df*=153; *p*<0.001. PA revealed that the best solution would be a 3-factor structure that explains 53.54% of the variance cumulatively. Table 2 shows factor loadings of items and percentage of explained variance per factor.

**Table 2.** Factor Loadings and Explained Variance of the PAWEC.

| Item | Factor loading |  |  |
| --- | --- | --- | --- |
|  | Factor 1 | Factor 2 | Factor 3 |
| 1. Do you feel good by practicing exercise during this period of quarantine? | 0.93 | -0.08 | -0.04 |
| 7. Do you feel happy by practicing exercise during the period of social isolation? | 0.91 | -0.06 | -0.04 |
| 6. Do you feel more disposed by practicing exercise during this period of quarantine? | 0.84 | 0.06 | 0.02 |
| 2. Do you feel well the days you practice exercise during the period of social isolation? | 0.81 | -0.04 | 0.06 |
| 10. Do you feel bad whenever you do not exercise during quarantine? | -0.51 | -0.14 | 0.16 |
| 16. Do you feel more indisposed whenever you do not exercise during this period of quarantine? | -0.40 | 0.03 | 0.19 |
| 9. Do you believe it is important to exercise during this period of quarantine? | 0.03 | 0.67 | 0.27 |
| 15. Do you believe being necessary to exercise during quarantine? | 0.04 | 0.66 | 0.29 |
| 18. Do you believe that exercising during quarantine is unimportant? | -0.11 | -0.51 | -0.13 |
| 4. Do you feel your day gets better whenever you exercise during this period of quarantine? | 0.03 | 0.45 | 0.10 |
| 11. Do you feel your day gets worse whenever you do not exercise during this period of quarantine? | 0.08 | -0.35 | -0.14 |
| 8. Do you feel more motivated to exercise during this period of quarantine? | 0.12 | 0.24 | 0.53 |
| 3. Does your mood improve whenever you exercise during the period of social isolation? | 0.02 | 0.09 | 0.51 |
| 17. Do you feel unmotivated to exercise during this period of quarantine? | -0.14 | 0.03 | -0.47 |
| 5. Do you feel less anxious while practicing exercise during the period of social isolation? | 0.25 | 0.11 | 0.43 |
| 14. Do you feel more anxious the days you do not practice exercise during the period of social isolation? | -0.02 | -0.16 | -0.42 |
| 13. Do you feel sad whenever you do not practice exercise during quarantine? | -0.11 | -0.04 | -0.37 |
| 12. Does your mood worsen whenever you do not practice exercise during quarantine? | -0.22 | 0.03 | -0.31 |
| Explained Variance (%) | 23.99% | 17.13% | 14.42% |

Next, three dimensions were named based on the content of items loaded in the same factor: exercise effects (positive or negative affect of exercise for people in quarantine), cognition (cognitive variables which entail understanding reasons and importance of exercise during confinement) and mood (motivational and emotional aspects involved in exercising during social isolation). Reliability of the entire PAWEC was α=0.84. Separately, the factor *exercise effects* presented α=0.77, the factor *cognition* showed α=0.71 and the factor *mood* had α=0.82.

After reassuring the validity and reliability of PAWEC, the first LMR computed was the Positive Affect score of PANAS as the dependent variable. The model was significant for *F*(5,30)=30.850; *p*<0.001; *f*^2^=0.06 with 19% of the variance explained according to the *r*^2^=0.19. Regarding the Negative Affect factor of PANAS, the model was also significant for *F*(7,28)=35.498; *p*<0.001; *f*^2^=0.08 and coefficient of determination *r*^2^=0.22, which suggests that the variance of NA explained by independent variables was 22%. Table 3 provides statistical coefficients of the two LMR.

**Table 3.** Results of the LMR having PA and NA scores of PANAS separately as dependent variables

| Variable | LMR Statistics |  |  |
| --- | --- | --- | --- |
| | $\beta$ | *t*-test | *p*-value |
| *Positive Affect score (PANAS)* |  |  |  |
| (Intercept) | 32.537 | 21.180 | <0.001 |
| PAWEC *mood* score | 2.017 | 4.409 | <0.050 |
| Use of media source during quarantine | -1.629 | -4.464 | <0.050 |
| Frequency of exercise before isolaton | 1.323 | 3.439 | <0.050 |
| PAWEC *effects of exercise* score | 0.380 | 2.200 | <0.050 |
| *Negative Affect score (PANAS)* |  |  |  |
| (Intercept) | 20.121 | 14.398 | <0.001 |
| PAWEC *effects of exercise* score | -4.028 | -11.958 | <0.001 |
| PAWEC *mood* score | -2.923 | -3.742 | <0.001 |
| Number of days in quarantine | 2.170 | 3.236 | <0.001 |
| Increase in exercise frequency | 1.031 | 2.837 | <0.005 |
| Gender | 0.956 | 2.703 | <0.050 |
| Use of media source during quarantine | 0.474 | 2.475 | <0.050 |

## Discussion

This study aimed to understand the relationship between physical activity and SWB among individuals in the social isolation period due to COVID-19. Considering the presented results, the practice of physical activity during the quarantine was associate to SWB in the aspects regarding mood, effects of exercise, gender, number of days in social isolation, the use of media resources and the frequency of physical activity before and during the quarantine.

Although the main goal of this research was to understand the relationship between aspects of exercise and SWB, an instrument was developed for measuring attitude of individuals toward exercise during quarantine. The PAWEC was a scale created for this study and factorial analysis pointed to a three-factor model as the best framework to explain the latent structures of physical activity among people in confinement situations. Exploratory factor analysis were conducted according to Baglin’s (2014) suggestions and revealed good sample adequacy and capacity for factorial rotation, which was required since the instrument showed a multidimensional structure. The first PAWEC’s dimension was mood (i.e., either positive or negative effects of exercise for people in quarantine, but not necessarily after exercising, more of an overall feeling such as feeling less anxious or feeling more motivated to work out during the quarantine). The second PAWEC’s factor found in this EFA was named exercise effects (i.e., motivational and emotional positive aspects involved in exercising during social isolation, such as feeling happy or feeling well disposed after exercising). The last of the PAWECs dimensions was cognition (i.e., understanding reasons and importance of exercise during confinement such as believing being necessary to exercise and understanding to be important to practice physical activity). All factors separately and the scale as a whole showed Cronbach’s alpha above 0.70, which was considered enough to assume good internal consistency. The instrument, then, proved to be reliable and well structured.

Results suggested that the exercise effects dimension of the PAWEC was correlated with both negative and positive affect subscale of the PANAS. The more the person feels that physical activity during confinement has a clear effect in their feelings, the greater the association to both dimensions of the SWB. It means that a person who does not enjoy exercising will probably not feel neither positive nor negative aspects of the SWB. The neurophysiological, social and interactional benefits of regular physical activity are already known (Abrantes et al., 2012; Buffart et al., 2012; Edenfield & Saeed, 2012). These benefits are also maintained during a social isolation situation, as experienced in the pandemic period of Covid-19, especially when we take into consideration the mood factor. However, SWB as measured by PANAS is only associated whenever the person actually experiences either the subjective feeling of happiness and physical disposition after exercising or any negative effects of exercising. For those people who do not feel any changes after exercising, the SWB will remain unaltered.

Another dimension of the PAWEC, exercise effects, also showed significant relationship with both positive and negative affects of the SWB. The literature has shown previously that exercising leads to improving well-being and mental health outcomes (Samulski & Noce, 2000; Stuart & Nanette, 2007; Panza et al., 2019). Accordingly, feeling better during the quarantine due to Covid-19 is something that can be linked to exercise routine, since it prevents depression, anxiety and stress levels to increase (Filgueiras & Stults-Kolehmainen, 2020). So, these findings corroborate with previous literature that suggests that exercising frequently helps the positive aspects of the SWB.

Even though there are studies that highlight the relevance of individuals’ awareness of exercise benefits to engage and endure in its practice and to improve SWB (Santos et al., 2009), the present evidence suggests that being aware of the role of exercise does not lead to either improvement or decrease of SWB among people in quarantine due to Covid-19.

Therefore, just saying that exercising during quarantine is important does not have any effect on people’s well-being. Practitioners must rely on preparing exercise routines that are joyful and ludic instead of worrying on explaining individuals the relevance of exercising, at least in situations of social isolation.

Beyond attitude towards exercise of participants, more objective variables were collected in the present study. Gender correlated to negative affect of the SWB. Due to the coding (i.e., men was coded as #1 and women coded as #2) the positive linear relationship between negative affect and gender means that women showed higher negative affect of the SWB when compared to men. This evidence is supported by previous findings that women suffer more anxiety, stress and depression when compared to men during quarantine due to Covid-19 (Filgueiras & Stults-Kolehmainen, 2020), so it makes sense that women feel more discomfort in those conditions.

Another evidence that could be expected to be found was the association between number of days in quarantine and negative affect. The longer the quarantine, the worse are the feelings regarding SWB. Those findings corroborate with evidence from other studies of mental health among people in quarantine due to other epidemic conditions (Bai et al., 2004; Hawryluck et al., 2004; Lee et al., 2005). A long time of social isolation and confinement increases the risk for mental health symptoms, thus, quarantine must be a temporary rather than a permanent resource for avoiding contamination.

Participants answered questions regarding their frequency and type of physical activity. Although type of exercise did not differ in the present results, two exercise-related variables were significantly associated to SWB: frequency of exercise before the quarantine and increase of frequency of exercise during quarantine. The literature indicates that three to five days a week of moderate exercise increases positive affect and decreases negative affect of the SWB (Stuart & Nanette, 2007; Buckworth & Dishman, 2002). In fact, those participants who reported to exercise before the quarantine and kept exercising showed higher association with positive affect of the SWB than those who did not engage in physical activity routines before the quarantine. On the opposite side of the SWB, increasing in the frequency of exercise was positively associated to negative affect, which means that those who reported more exercise during quarantine felt worse than those who kept their routines. Stuart and Nanette (2007) suggest that sudden increases in exercise routine as well as exhaustive exercises above five days a week may lead to less SWB. What the findings from this study support is that balance is pivotal regarding exercise routine, so, both sedentary and exaggerate exercise behaviours may increase negative affect of one’s SWB. In this sense, a physical activity program to introduce more benefits must be individualized and developed by a physical education teacher who has knowledge of these training principles in order to achieve maximum exercise performance and protect the body against harmful excesses, such as injuries (Bessa, Silva, Carrijo, & Oliveira, 2013). Thus, people who included physical activity practice only after quarantine may not have taken these principles into account and this will generate feelings of discomfort.

The study by Filgueiras and Stults-Kolehmainen (2020) on mental health during quarantine due to Covid-19 showed that sudden changes in habits during the isolation period increases the risks for depression, anxiety and acute stress. This result corroborates with these found in the present study. Quarantine provides people the sense of that starting regular physical activity could contribute to their physical and mental health. However, it seems clear that this sudden inclusion of an exercise routine, and often without prior planning, can contribute to a greater sense of subjective malaise.

An important aspect highlighted by Becker Junior (2000) is that in order to observe the effects of exercise and sport on the emotional area, it is necessary to respect the practice time, varying between four and twenty weeks. The intensity with which physical exercise is performed is also a factor, which interferes with the type of emotional response observed. The more frequent the exercise practice, the less presence of psychological disorders and many stressors tend to reduce their strength as the exercise habit increases (Krause et al., 1993). Thus, the inclusion of physical activity only in the quarantine period is not associated with greater subjective well-being. This result points out that the inclusion of healthier habits in the social isolation period should not start with an increase in physical activity practice.

The sudden introduction of physical activity and the lack of individualized planning lead to another issue found in this research: the relationship between SWB and the adoption of social media and other internet resources to build exercise routines. Those who used technological yet impersonalized applications (i.e., YouTube, Instagram and other mobile apps) to conduct their practices of physical activity had a decrease in the positive affect of the SWB and an increase of negative affect of the SWB. These classes of training designs do not take into account the reality of each individual, they are generalists, and, therefore only meet the demands of a specific group of people (Bessa et al, 2013). Although it could be expected that those resources actually are beneficial or, at least, unassociated to SWB, in fact they decrease positive affect and increase negative affect, which makes them not recommended for people in quarantine due to Covid-19.

Based on the results described herein, physical activity practice during quarantine can contribute to a greater SWB for people in quarantine due to Covid-19, but only those who were already in the habit of exercising. Including a new habit, which requires guidance and care during isolation, can induce increase of negative affect. In addition, indiscriminate use of media resources to initiate practice brings greater sense of discomfort. It is important that the practice of physical activity in a period of social isolation is designed to be closer to previous habits; including an obligation to exercise during this period when this was not a reality for the person can contribute to an increase in malaise. Maybe other strategies such as investing in a balanced diet and regular feeding habits (Filgueiras & Stults-Kolehmainen, 2020), getting involved with artistic activities can be better ways to increase well-being.

## Data Availability

Data is available upon request to one of the authors. In case of any inquiry, please contact Dr. Filgueiras

## Acknowledgements

Author collaborated equally to the study and manuscript. Authors thank the support of the Coordenacao de Aperfeicoamento de Pessoal de Nivel Superior (CAPES) and the Fundacao Carlos Chagas de Amparo a Pesquisa do Estado do Rio de Janeiro (FAPERJ) to conduct this study.

